# Single-Cell Transcriptome-Wide Mendelian Randomization and Colocalization Analysis Reveals Immune Cell-Specific Mechanisms in Alzheimer’s Disease

**DOI:** 10.64898/2026.11.03.26367234

**Authors:** Youwei Si, Youwen Si

## Abstract

**Objective:** To integrate single-cell expression quantitative trait locus (sc-eQTL), Alzheimer’s disease (AD) genome-wide association study (GWAS), and Bayesian colocalization analyses, in order to systematically identify candidate causal genes and potential immune mechanisms that specifically regulate AD risk in peripheral immune cells.

**Methods:** Using the cis-eQTLs of 14 peripheral blood immune cell types from the OneK1K single-cell eQTL dataset as exposure data and the AD GWAS (GCST90565439) as outcome data, a two-sample Mendelian randomization (MR) analysis was performed. Weak instrumental variables were removed using F-statistics, and approximately independent genetic instruments were obtained by linkage disequilibrium pruning. For significant gene–cell type associations, compareCluster functional enrichment, Bayesian colocalization, and phenome-wide association study (PheWAS) were further performed, and the 1M-scBloodNL dataset was used for replication validation.

**Results:** The MR analysis identified 187 gene–cell type associations significantly associated with AD risk, involving 161 candidate causal genes. Among them, 141 genes showed significant associations in only a single immune cell type, suggesting that AD-related immune genetic regulation has marked cell type specificity. Functional enrichment analysis showed that the significant genes in different immune cell types were enriched in biological processes including MHC class II antigen presentation, phagocytosis, membrane invagination, and positive regulation of T cell activation. Colocalization analysis indicated a strong shared genetic signal between TNF expression in CD4+SOX4+ T cells and AD risk (PP.H4 = 0.833). In the 1M-scBloodNL replication analysis, several candidate genes showed consistent direction and nominal significance. The PheWAS results suggested that the significant phenotypic associations of the TNF instrumental variable were mainly concentrated in immune- and hematology-related traits, with no genome-wide significant neurological phenotypic associations observed.

**Conclusion:** This study suggests that single-cell eQTL-MR combined with colocalization analysis can be used to dissect the cell type-specific genetic regulatory mechanisms of peripheral immunity in AD. The genetically predicted expression of TNF in CD4+SOX4+ T cells may represent an important immune-related signal for AD risk, but further validation in independent populations and functional experiments is still required.

## 1 Introduction

Alzheimer’s disease (AD) is the most common type of dementia and an important neurodegenerative disease that leads to cognitive decline and reduced quality of life in the elderly. With the intensification of population aging, the number of AD patients continues to increase, and AD has become a major challenge in the field of global public health.

Epidemiological data show that the global burden of AD and related dementias is substantial; the number of patients with AD and related dementias in China has also shown a continuous upward trend, and the prevalence in women is markedly higher than in men [1, 2].

The pathological features of AD mainly include amyloid deposition, abnormal tau protein phosphorylation, synaptic damage, and chronic neuroinflammation. In recent years, neuroimmunological studies have suggested that the peripheral immune system may be involved in the development and progression of AD. Peripheral immune cells such as T cells, B cells, monocytes/macrophages, and neutrophils can influence AD pathological processes through mechanisms including the release of inflammatory factors, immune cell migration, meningeal and perivascular immune surveillance, and central–peripheral immune crosstalk [3-8]. However, which gene regulatory events in different immune cell types have potential causal relationships with AD risk remains incompletely understood.

GWAS have identified multiple genetic loci associated with AD risk and have suggested that pathways such as lipid metabolism, endocytosis/phagocytosis, microglial function, and immune regulation play important roles in AD [9-13]. However, GWAS signals are usually located in non-coding regions and are affected by linkage disequilibrium and the complexity of regulatory networks, making it difficult to directly pinpoint the specific effector genes and their cell types of action. Transcriptome-wide association studies (TWAS), summary-data-based MR (SMR), protein quantitative trait locus MR, and drug target prioritization analyses have been used to screen candidate causal genes and potential therapeutic targets for AD [14-19], but most studies rely on bulk tissue, mixed cell populations, or protein-level QTL data, which may mask key cell type-specific effects.

Single-cell eQTL data provide new opportunities for dissecting cell type-specific genetic regulation [20-23]. By combining single-cell eQTL, AD GWAS, MR, and colocalization analyses, candidate causal genes whose genetically predicted expression is associated with AD risk can be identified at cell type resolution. In this study, we integrated peripheral blood immune cell sc-eQTL data from OneK1K and 1M-scBloodNL to systematically evaluate the potential causal effects of gene expression in 14 immune cell types on AD risk, and further explored their biological significance and potential pleiotropy through functional enrichment, colocalization, and PheWAS analyses.

## 2 Materials and Methods

### 2.1 Study Design and Ethical Statement

This study was designed in accordance with the STROBE-MR reporting guidelines [24]. This study is a secondary analysis using existing data, and only uses non-personally identifiable single-cell eQTL, GWAS, and PheWAS summary statistics published by the source studies; it does not involve the recruitment of new participants, the collection of human samples, interventions, or the processing of personally identifiable data. The recruitment of participants, informed consent, and ethical approval in the original studies were the responsibility of the respective original studies; this study did not conduct new participant research, and therefore did not apply for additional ethical approval, and complied with the data use conditions of the databases used. The overall study workflow included: screening of instrumental variables from immune cell sc-eQTL, two-sample MR analysis, functional enrichment analysis, colocalization analysis, replication validation using an independent sc-eQTL dataset, and PheWAS analysis.

### 2.2 Exposure Data and Instrumental Variable Selection

The exposure data were derived from the OneK1K single-cell eQTL resource [20]. This resource was constructed from 1,267,758 peripheral blood mononuclear cells (PBMCs) from 982 healthy donors and reported 26,597 independent cis-eQTLs and 990 trans-eQTLs across 14 immune cell types. The included cell types comprised CD4 naive/central memory T cells, CD4 effector/memory T cells, CD4 SOX4+ T cells, CD8 naive/central memory T cells, CD8 effector/memory T cells, CD8 S100B+ T cells, classical monocytes, non-classical monocytes, dendritic cells, natural killer cells, natural killer recruiting cells, immature B cells, memory B cells, and plasma cells.

This study directly adopted the cis-eQTLs defined in the original OneK1K dataset and did not redefine cis-eQTLs according to a ±1 Mb or ±100 kb genomic window. In the discovery-phase MR analysis, the threshold for screening candidate instrumental variables was P ≤ 1×10^−5^, and the strength of instrumental variables was assessed using the F-statistic (F = β^2^/SE^2^), with weak instrumental variables with F ≤ 10 removed [25]. To obtain approximately independent instrumental variables, LD pruning was performed using the clump_data() function in the TwoSampleMR package, with parameters set to a window of 10,000 kb, r^2^ = 0.001, and a clump P threshold of 1 [26].

The replication analysis used single-cell cis-eQTL data under unstimulated conditions from the 1M-scBloodNL dataset [21]. This dataset contains 928,275 PBMCs from 120 individuals and covers 6 major immune cell types: B cells, CD4+ T cells, CD8+ T cells, natural killer cells, monocytes, and dendritic cells. In the replication analysis, the 14 cell types of OneK1K were mapped to the corresponding 6 cell types of 1M-scBloodNL. For each target gene, the lead cis-eQTL with the smallest eQTL P value in the corresponding cell type was selected as the instrumental variable; given the smaller sample size of the replication cohort, no minimum eQTL P value threshold was set.

### 2.3 Outcome Data

The AD outcome data were obtained from GCST90565439 in the GWAS Catalog [27, 28]. This GWAS included 99,796 individuals of European ancestry, comprising 36,659 AD cases and 63,137 controls. To reduce the influence of population structure differences on the MR results, both the exposure and outcome data in this study were restricted to European ancestry-related data.

### 2.4 sc-eQTL Mendelian Randomization Analysis

Two-sample MR was used to assess the association between immune cell-specific genetically predicted gene expression and AD risk. Only exposure SNPs that could be directly matched in the AD GWAS summary statistics were retained; proxy SNPs were not used. The harmonise_data() function in TwoSampleMR was used for effect allele harmonization; in the discovery phase, action = 2 was used to remove palindromic SNPs whose direction could not be determined [26].

For genes containing only 1 instrumental variable, the Wald ratio method was used to estimate the effect; for genes containing 2 or more instrumental variables after LD pruning, the inverse-variance weighted (IVW) method was used as the primary MR method. When the number of instrumental variables was sufficient, sensitivity analyses were further performed using the weighted median method, MR-Egger regression, the weighted mode method, heterogeneity testing, and leave-one-out analysis [29-31]. MR results were expressed as odds ratios (ORs) with 95% confidence intervals (CIs), and the Benjamini-Hochberg false discovery rate (FDR) method was used for multiple-testing correction [32]. FDR < 0.05 was considered statistically significant.

### 2.5 Functional Enrichment Analysis

Cell type-specific functional enrichment analysis was performed using the compareCluster() function in the clusterProfiler package [33]. The MR genes that were significant in the primary IVW analysis were grouped by immune cell type, and gene symbols were converted to Entrez IDs using org.Hs.eg.db. Cell types with fewer than 5 mapped genes were excluded. GO biological process enrichment analysis was performed, and multiple-testing correction was performed using the Benjamini-Hochberg method; terms with adjusted P < 0.05 were considered significant [32]. Enrichment results were visualized using dot plots.

### 2.6 Colocalization Analysis

To determine whether the MR results were driven by gene expression and AD risk sharing the same causal genetic variant, Bayesian colocalization analysis was performed using the coloc R package [34, 35]. This method evaluates 5 mutually exclusive hypotheses: H0 indicates no association with either trait; H1 indicates association with gene expression only; H2 indicates association with AD risk only; H3 indicates association with both traits but driven by different causal variants; and H4 indicates that both traits are driven by the same shared causal variant. In this study, PP.H4 > 0.75 was defined as strong colocalization evidence, and 0.50 < PP.H4 ≤ 0.75 was defined as moderate colocalization evidence [36].

### 2.7 PheWAS Analysis

To assess the broad phenotypic associations and potential pleiotropy of the prioritized candidate genes and key instrumental variables, gene-level and SNP-level PheWAS analyses were performed. The gene-level analysis used the AstraZeneca PheWAS Portal [37]. For the TNF instrumental variable rs3130043 in CD4 SOX4 cells, the IEU OpenGWAS database and the FinnGen R12 database were queried separately [38]. The genome-wide significance threshold was set at P < 5×10^−8^, and significant associations were classified into immune-related, neurological, hematological, metabolic-related, and other categories according to the phenotype description.

## 3 Results

### 3.1 Identification of AD-Related Candidate Causal Genes

Based on cis-eQTLs from 14 immune cell types, the MR analysis identified a total of 187 gene– cell type associations significantly associated with AD risk (FDR < 0.05), involving 161 unique genes (Figure 1, Supplementary Table 1). Among them, 141 genes (87.6%) were significantly associated with AD risk in only a single immune cell type, accounting for 75.4% (141/187) of all significant gene–cell type associations. This result suggests that AD-related immune genetic regulation has marked cell type specificity and is more likely to act through narrow-range transcriptional regulatory programs in specific immune cell lineages, rather than through broadly shared changes in immune expression.

**Figure 1.**
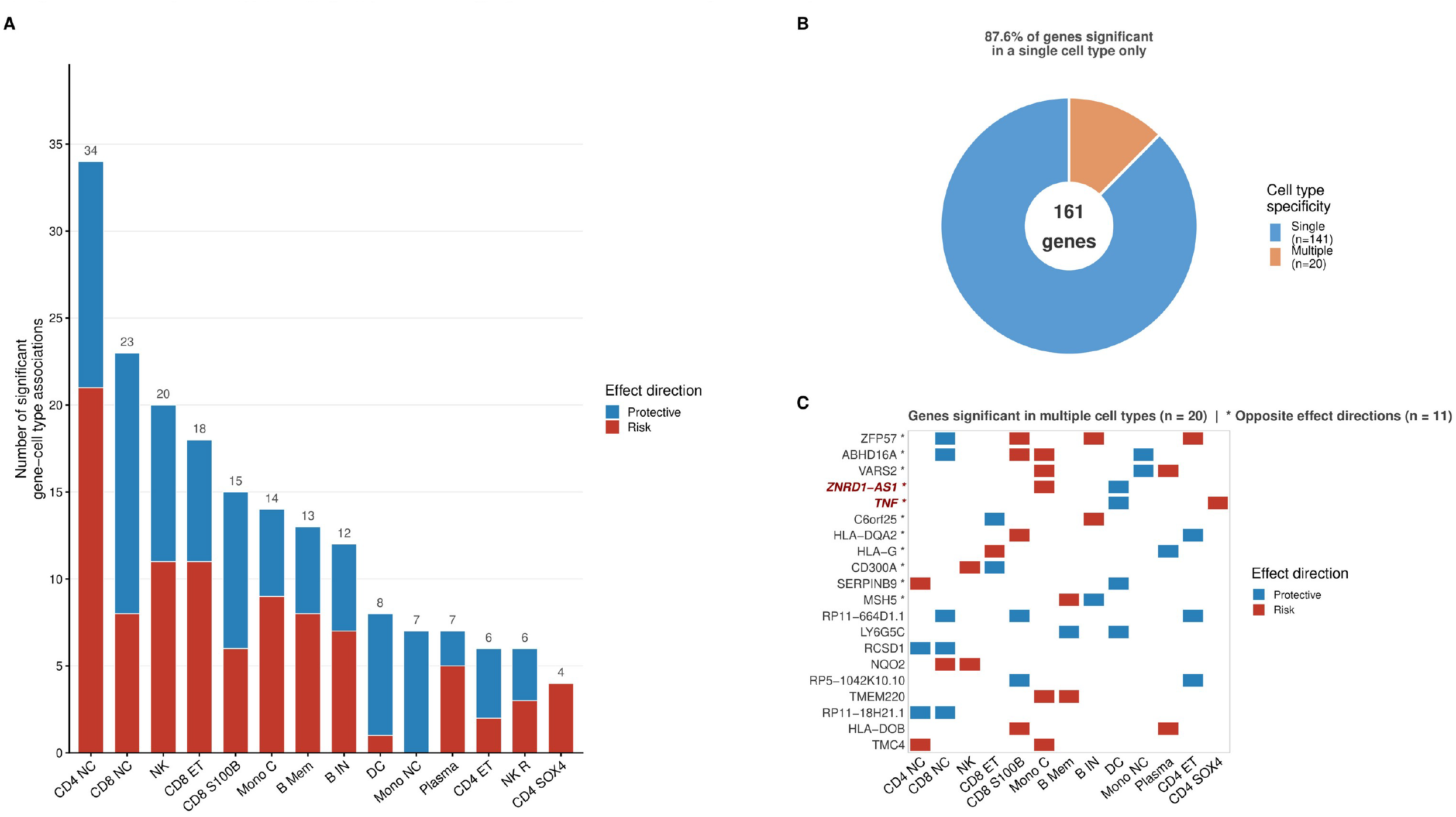
Cell type specificity of MR associations between immune cell gene expression and AD risk. A. Number of FDR-significant gene–cell type associations in each immune cell type. Red/blue indicate risk/protective effect directions, respectively. B. Proportion of single-cell-type-specific genes (blue, n = 141, 87.6%) versus multi-cell-type genes (orange, n = 20) among the 161 significant genes. C. Heatmap of effect directions for the 20 multi-cell-type genes. Asterisks (*) mark genes with opposite effect directions across cell types (n = 11); TNF and ZNRD1-AS1 are highlighted in bold red.

Among the 20 genes significant across multiple cell types, 11 genes showed opposite effect directions in different cell types. For example, elevated genetically predicted TNF expression in CD4 SOX4 cells was associated with increased AD risk, whereas in dendritic cells it showed a protective direction; ZNRD1-AS1 showed an increased-risk direction in classical monocytes and a protective direction in dendritic cells. These results further emphasize that using only bulk tissue or mixed cell population eQTL data may mask or even cancel out cell-specific genetic effects with opposite directions.

### 3.2 Cell Type-Specific Functional Enrichment Results

To explore the biological functions of AD-related MR genes in different immune cells, compareCluster was used to perform GO biological process enrichment analysis stratified by cell type. The results showed that significant genes in different immune cell types were enriched in different immune-related biological processes (Figure 2). Significant genes in CD8 S100B+ T cells were mainly enriched in MHC class II antigen presentation-related processes, with representative genes including HLA-DOA, HLA-DQA2, and HLA-DOB. In CD4 naive/central memory T cells, significant genes were enriched in processes such as phagocytosis and membrane invagination, with representative genes including ABCA7, ELMO1, and ITGA2. Plasma cell-related genes were enriched in positive regulation of T cell activation, with representative genes including AIF1, HLA-G, and HLA-DOB.

**Figure 2.**
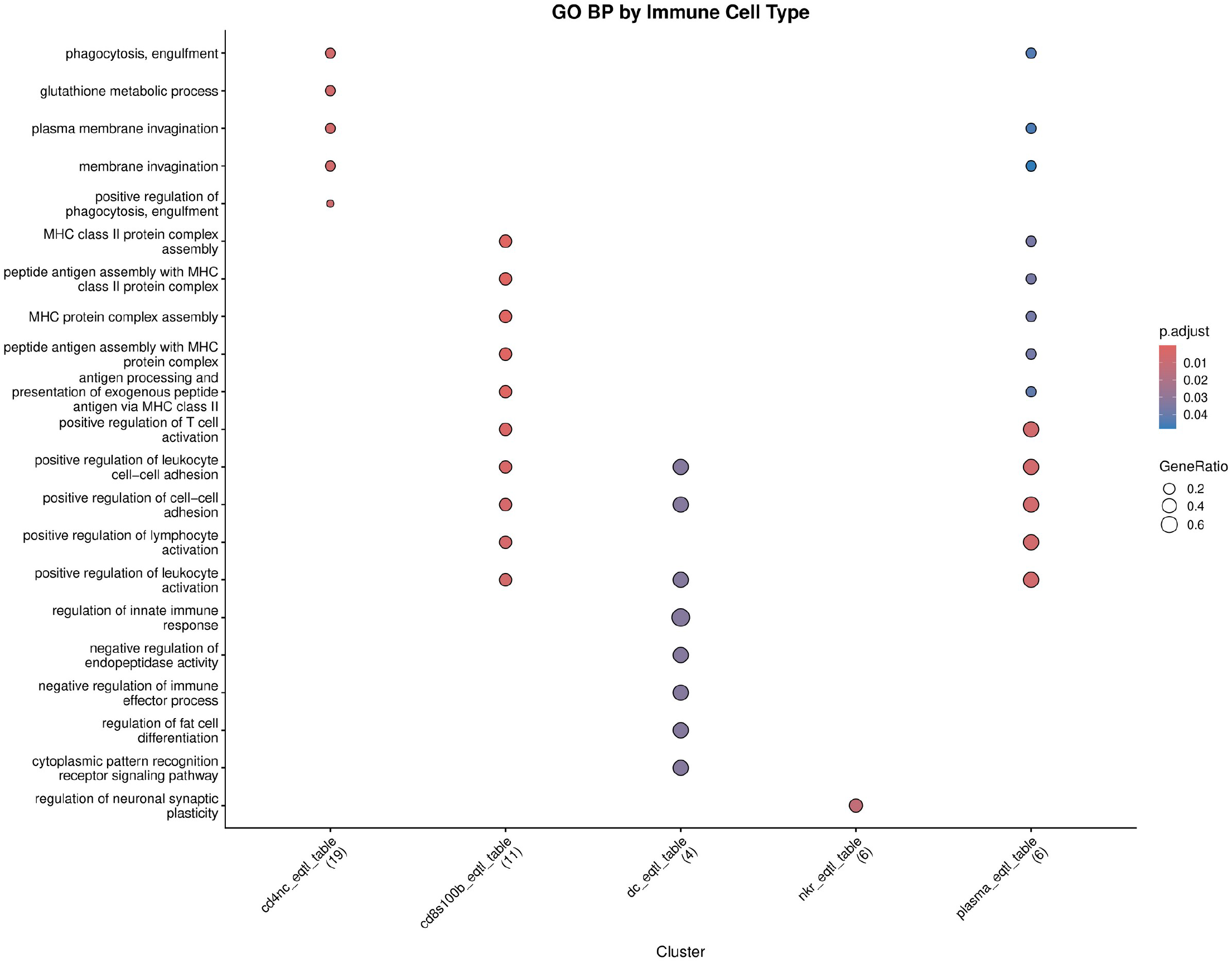
GO-BP enrichment analysis of AD-related genes in different immune cell types. compareCluster was used to perform GO biological process enrichment analysis of significant MR genes by immune cell type. The x-axis represents immune cell types, and the y-axis represents enriched terms; bubble size represents GeneRatio, and color represents adjusted P value. The results show that AD-related genes exhibit cell type-specific enrichment in different immune cells, mainly involving immune-related processes such as MHC class II antigen processing and presentation, phagocytosis, membrane invagination, T cell activation, and leukocyte adhesion.

These results suggest that AD-related immune genetic signals are not concentrated in a single immune pathway, but rather involve antigen presentation, phagocytic uptake, membrane dynamics, and lymphocyte costimulation in different immune cell types.

### 3.3 Colocalization Analysis Prioritizes Immune Cell-Specific AD-Related eGenes

To determine whether the significant MR associations were driven by gene expression and AD risk sharing the same genetic variant, Bayesian colocalization analysis was performed on FDR-significant gene–cell type combinations. The strongest colocalization evidence was found at the TNF locus in CD4+SOX4+ T cells. This locus included a total of 1,596 overlapping SNPs, with a PP.H4 of 0.833, suggesting that TNF expression and AD risk are likely driven by the same shared genetic signal (Figure 3).

**Figure 3.**
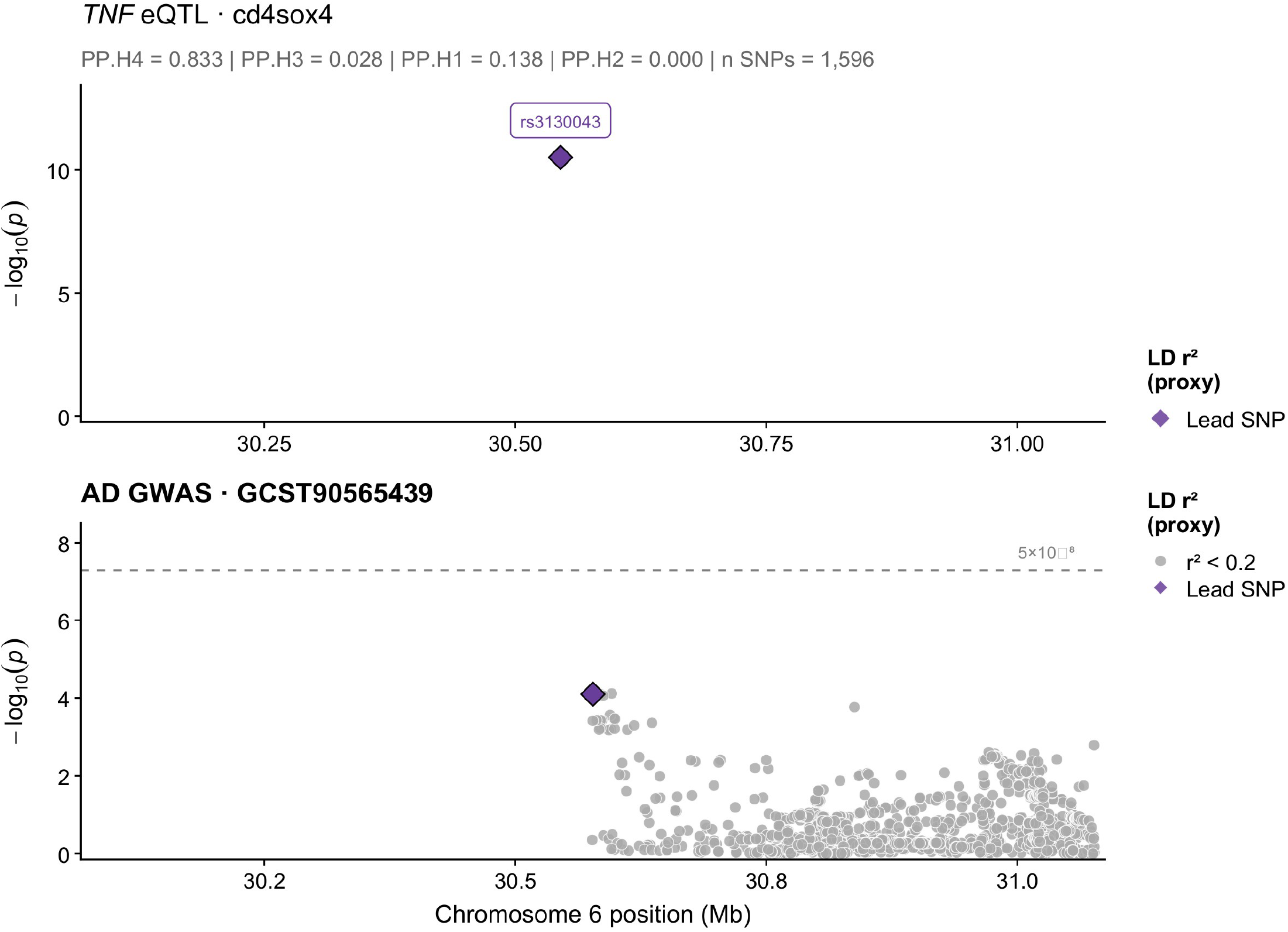
Bayesian colocalization analysis of the TNF eQTL signal and the Alzheimer’s disease risk locus. Regional association plots of SNPs within a ±500 kb window centered on the lead variant rs3130043 (chromosome 6). The top panel shows the strength of association of this variant with TNF expression in CD4^+^SOX4^+^ T cells (OneK1K, n = 982); the bottom panel shows the strength of association of SNPs in the same window with AD risk (GCST90565439, N = 99,796). The purple diamond (◆) indicates the lead variant rs3130043, and the horizontal dashed line indicates the genome-wide significance threshold (p = 5×10^−8^). Colocalization analysis used the Bayesian coloc.abf framework, PP.H4 = 0.833, suggesting that the two traits share the same causal genetic variant at this locus (number of overlapping SNPs = 1,596).

The MR analysis showed that elevated genetically predicted TNF expression in CD4 SOX4 cells was significantly associated with increased AD risk (OR = 1.177, 95% CI: 1.138–1.217, P = 1.72×10^−21^, FDR = 2.94×10^−19^). This MR instrumental variable consisted of 10 independent SNPs, with a mean F-statistic of 57.0, indicating high instrumental variable strength. The consistency between the MR and colocalization analysis results supports that TNF expression in CD4 SOX4 T cells may be a high-priority immune genetic signal associated with AD risk.

### 3.4 Replication Validation in an Independent sc-eQTL Dataset

To assess the reproducibility of the main findings, replication analysis was further performed using the 1M-scBloodNL dataset. After mapping the 14 cell types of OneK1K to the 6 immune cell types of 1M-scBloodNL, a total of 171 gene–cell type combinations could be preliminarily evaluated; among them, 40 combinations yielded replication results that allowed comparison of MR effect directions between the two datasets (Figure 4).

**Figure 4.**
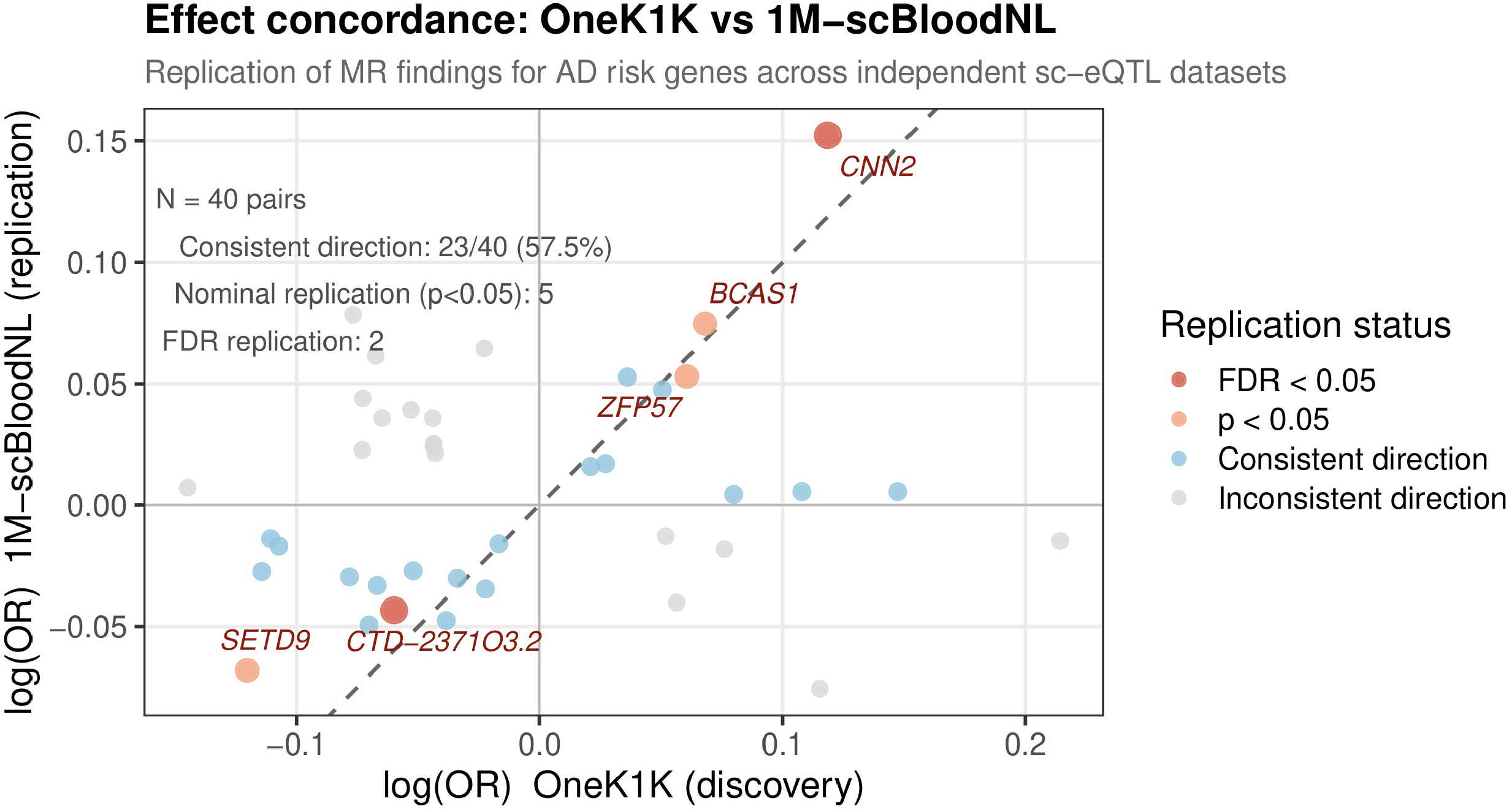
Scatter plot of MR effect consistency between the OneK1K and 1M-scBloodNL datasets. The x-axis is the log(OR) value in the OneK1K discovery cohort, and the y-axis is the log(OR) value in the 1M-scBloodNL replication cohort. The dashed line is a reference line with a slope of 1, indicating completely consistent effect sizes between the two datasets. Each data point represents a gene–cell type combination (N = 40), and color indicates replication status: dark red, significant after FDR correction (FDR < 0.05); light red, nominally significant (p < 0.05); blue, consistent effect direction but not significant; gray, inconsistent effect direction. Italic labels indicate the 5 nominally significant replication genes. A total of 23/40 combinations (57.5%) had consistent effect directions, of which 5 reached nominal significance (p < 0.05), and CTD-2371O3.2 and CNN2 remained significant after FDR correction.

Among these 40 combinations, 23 showed effect directions consistent with the OneK1K discovery phase, with a direction consistency rate of 57.5%. Among them, ZFP57 (CD4T, OR = 1.054), CTD-2371O3.2 (CD8T, OR = 0.958), SETD9 (CD8T, OR = 0.934), BCAS1 (CD4T, OR = 1.078), and CNN2 (CD4T, OR = 1.164) simultaneously showed consistent effect direction and nominal significance (P < 0.05). Among them, CTD-2371O3.2 and CNN2 remained significant after FDR correction, representing relatively robust replication candidate genes.

### 3.5 PheWAS Analysis of Prioritized Candidate Genes and Instrumental Variables

To assess the broad phenotypic associations and potential pleiotropy of the prioritized candidate signals, gene-level and SNP-level PheWAS analyses were performed for TNF. In the gene-level analysis of the AstraZeneca PheWAS Portal, no broad systemic phenotypic associations reaching genome-wide significance were observed for TNF (Figure 5).

**Figure 5.**
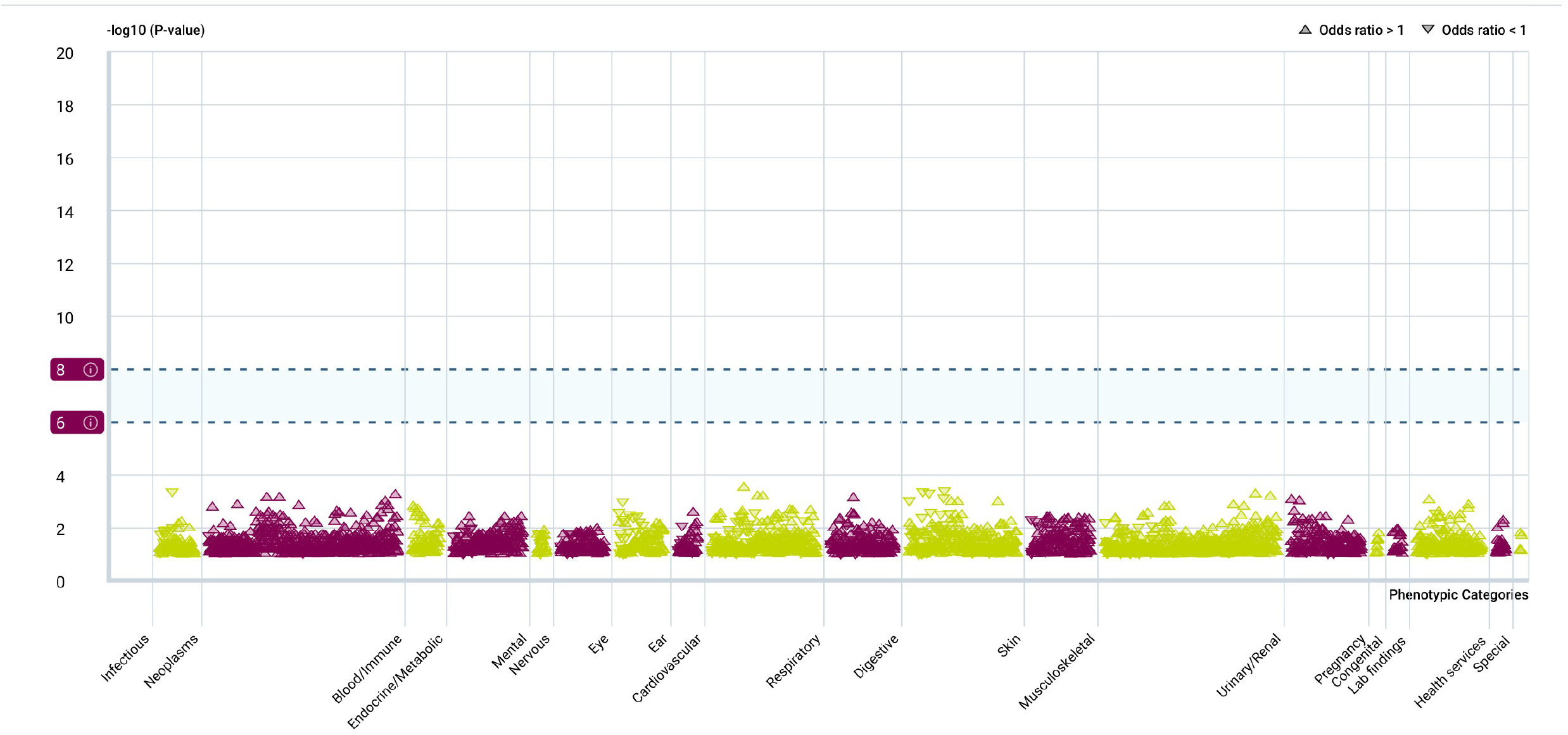
Gene-level PheWAS analysis results for TNF. Gene-level phenome-wide association analysis of TNF based on the AstraZeneca PheWAS Portal. The x-axis represents different phenotype categories, and the y-axis represents –log10(P value); triangles represent different phenotypic association signals, with direction distinguishing odds ratio > 1 and odds ratio < 1, and the dashed line represents the significance reference threshold. The results show that no broad systemic phenotypic associations reaching genome-wide significance were observed for TNF, suggesting that, as a prioritized candidate signal, it did not exhibit evident cross-system pleiotropy risk.

For the TNF instrumental variable rs3130043 in CD4 SOX4 cells, the IEU OpenGWAS analysis showed that a total of 105 phenotypic associations reached genome-wide significance (P < 5×10^−8^), mainly including immune-related traits (n = 37, such as eosinophil count, psoriasis, and celiac disease) and red blood cell-related indicators (n = 65, such as mean corpuscular volume and reticulocyte count) (Figure 6). In FinnGen R12, a total of 15 phenotypic associations reached genome-wide significance, all of which were immune-related diseases or phenotypes, including celiac disease, rheumatoid arthritis, ankylosing spondylitis, thyroiditis, and iridocyclitis (Figure 7). Notably, no neurological phenotypic associations reaching genome-wide significance were observed in either IEU OpenGWAS or FinnGen R12. This result suggests that the significant phenotypic spectrum of rs3130043 is mainly concentrated in immune- and hematology-related phenotypes; however, given that TNF is located in the MHC region, the pleiotropy of this instrumental variable should still be interpreted with caution.

**Figure 6.**
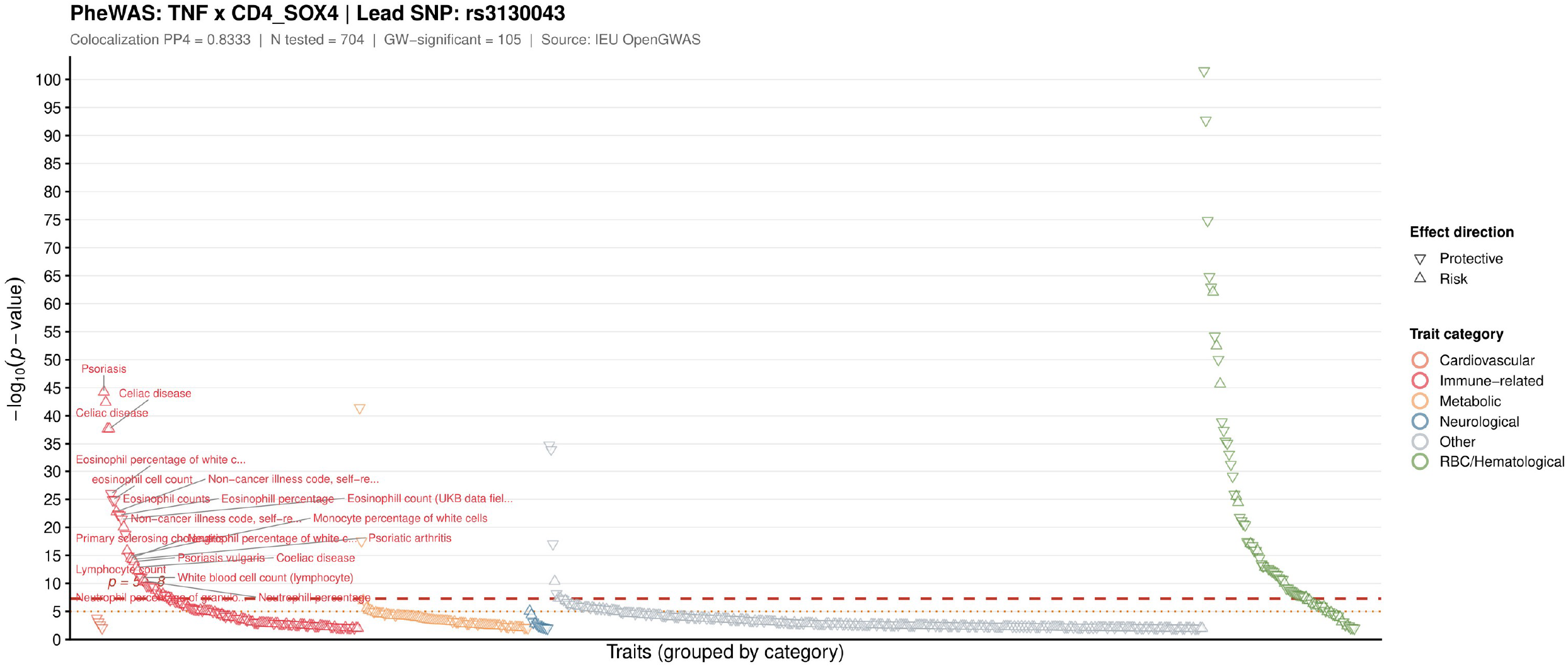
Phenome-wide association study (PheWAS) of the TNF genetic instrumental variable (rs3130043). The x-axis represents 704 tested traits grouped by category, and the y-axis represents –log_10_(p). The red dashed line indicates the genome-wide significance threshold (p = 5×10^−8^). Point color represents trait category, and triangle direction represents effect direction (up: risk; down: protective). Representative immune-related traits reaching genome-wide significance are labeled. A total of 105 traits reached genome-wide significance, predominantly immune-related traits (eosinophil count, psoriasis, celiac disease, etc.); no significant associations were observed for neurological traits. Data source: IEU OpenGWAS database. SNP: rs3130043 (TNF, CD4 SOX4 cells; colocalization PP4 = 0.833).

**Figure 7.**
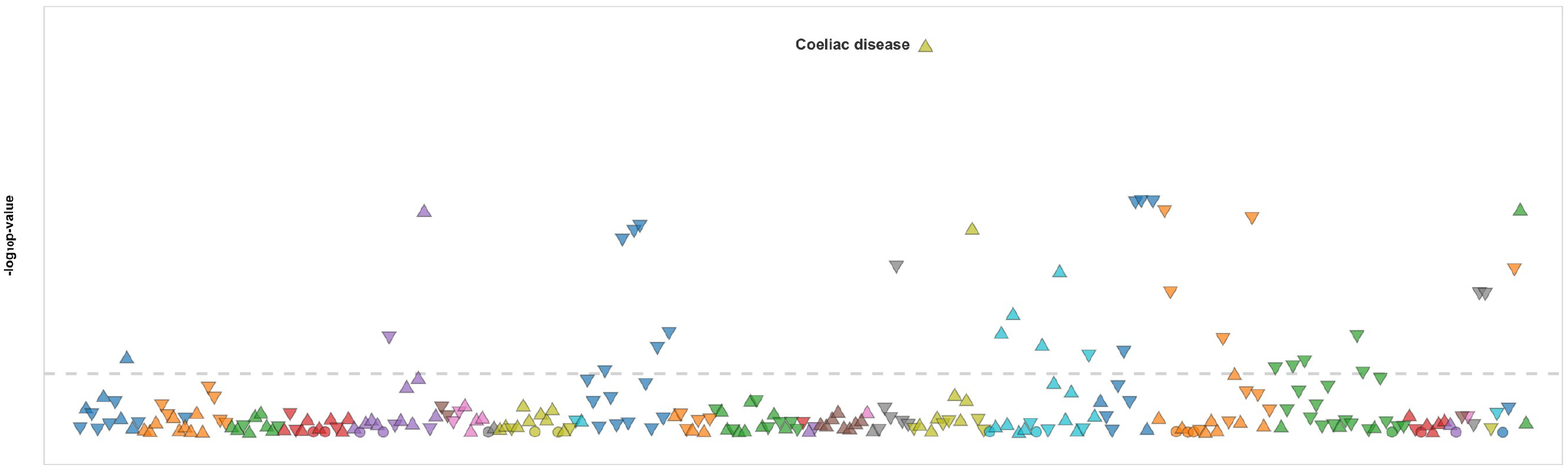
SNP-level PheWAS analysis results for the TNF instrumental variable rs3130043 in FinnGen R12. Phenome-wide association analysis of the TNF-related genetic instrumental variable rs3130043 (6-30577245-G-T) based on FinnGen Release 12. The y-axis represents –log10(P value), each point represents a FinnGen phenotype endpoint, and the dashed line indicates the genome-wide significance reference threshold. The results show that the significant associations of this variant were mainly concentrated in immune-related diseases, with celiac disease showing the strongest association signal; no evident significant associations with neurological diseases were observed, suggesting that the pleiotropy of this instrumental variable is mainly related to immune-related phenotypes.

## 4 Discussion

This study integrated sc-eQTL data from 14 peripheral blood immune cell types with a large-scale AD GWAS and, through two-sample MR, colocalization, and PheWAS analyses, systematically identified AD-related immune cell-specific genetic regulatory signals. The study identified a total of 187 significant gene–cell type associations, involving 161 candidate causal genes, the vast majority of which showed significant associations in only a single immune cell type. This result suggests that AD-related immune genetic mechanisms have marked cell type specificity, consistent with the cell type-dependent genetic regulatory features emphasized by recent single-cell eQTL studies [20-23].

An important finding of this study is that the same gene may exhibit opposite effect directions in different immune cell types. For example, TNF was associated with increased AD risk in CD4 SOX4 T cells but showed a protective direction in dendritic cells. Such opposite-direction cell-specific effects may explain why some genetic analyses based on bulk tissue, mixed blood cells, or plasma proteins have difficulty obtaining consistent results. Previous MR, TWAS, and drug target prioritization studies have identified multiple AD candidate targets [14-19], but the cell type context remains relatively insufficient. The advantage of single-cell eQTL-MR lies in its ability to localize genetic regulatory signals to more specific immune cell types, thereby improving the biological resolution of candidate target interpretation.

TNF was the most prominent candidate signal in this study. The MR analysis showed that elevated genetically predicted TNF expression in CD4 SOX4 T cells was significantly associated with increased AD risk, and the colocalization analysis further suggested that TNF expression and AD risk may share the same genetic signal. TNF is a classical pro-inflammatory cytokine involved in peripheral immune activation, amplification of inflammatory responses, and regulation of neuroinflammation [3]. Combined with previous studies on T cell infiltration and peripheral–central immune crosstalk in AD [4, 8], the results of this study suggest that TNF regulation in CD4 SOX4 T cells may be an important candidate mechanism linking peripheral immunity to AD genetic risk.

The functional enrichment results support that AD-related immune genetic signals involve multiple immune processes. The enrichment of MHC class II antigen presentation-related genes in CD8 S100B+ T cells suggests that antigen processing and presentation may be involved in the regulation of AD risk by adaptive immunity; the enrichment of genes such as ABCA7, ELMO1, and ITGA2 in CD4 naive/central memory T cells in phagocytosis and membrane invagination processes is consistent with the importance of the endocytosis/phagocytosis pathway in previous AD genetic studies [9-13]; the enrichment related to positive regulation of T cell activation in plasma cells suggests that B cell/plasma cell-related immune processes may be involved in the regulation of AD risk by affecting T cell responses.

This study also performed replication validation using the 1M-scBloodNL dataset [21]. Because this dataset has a smaller sample size and broader cell type classification than OneK1K, only some of the genes significant in the discovery phase could obtain corresponding lead eQTL instrumental variables. Nevertheless, some genes in the replication analysis still showed consistent direction and statistical support, suggesting that the discovery-phase results have a certain degree of reproducibility. Future larger-scale single-cell eQTL resources will help further validate these cell type-specific signals.

It should be noted that the PheWAS results cannot directly prove the clinical safety of the candidate targets. In this study, the significant phenotypic associations of rs3130043 were mainly concentrated in immune- and hematology-related traits, and no genome-wide significant neurological phenotypic associations were observed, which supports its specificity as an immune-related instrumental variable; however, TNF is located in the MHC region, which has complex linkage disequilibrium, polygenic regulation, and broad immune pleiotropy. Therefore, the therapeutic significance of the TNF–CD4 SOX4 T cell axis still needs to be further validated through more refined conditional colocalization, fine-mapping, protein-level evidence, and functional experiments [34, 35, 37, 38].

This study has several limitations. First, the discovery-phase sc-eQTL data and AD GWAS were mainly from populations of European ancestry, and the generalizability of the results to other populations still needs to be verified. Second, the eQTL data were derived from peripheral blood immune cells and cannot fully represent central nervous system-resident immune cells, especially the regulatory architecture of microglia. Third, MR analysis relies on the relevance, independence, and exclusion restriction assumptions of instrumental variables; although this study performed LD pruning, weak instrumental variable removal, sensitivity analysis, and colocalization analysis, horizontal pleiotropy cannot be completely excluded. Fourth, the LD structure of the MHC region is complex, and the interpretation of the TNF signal requires particular caution. Fifth, the replication dataset has a limited sample size and lower cell type resolution than the discovery dataset, which may reduce replication power.

## 5 Conclusion

This study shows that Mendelian randomization and colocalization analysis based on single-cell eQTL can reveal AD-related immune genetic mechanisms at cell type resolution. The study found that AD-related candidate causal genes have marked immune cell specificity, and prioritized TNF expression in CD4 SOX4 T cells as a high-priority immune genetic signal associated with AD risk. This study provides a methodological framework for dissecting AD immune mechanisms and discovering cell type-specific targets, but the related findings still require further genetic fine-mapping and functional experimental validation.

## Data Availability

This study performed secondary analysis using only publicly available summary statistics or public query results provided by existing studies. The data resources used and their access addresses are as follows (access date: November 4, 2026):
(1)OneK1K sc-eQTL (discovery phase): https://onek1k.org/.
(2)1M-scBloodNL sc-eQTL (replication phase): https://eqtlgen.org/sc/datasets/1m-scbloodnl.html.
(3)AD GWAS (GCST90565439, outcome data): https://www.ebi.ac.uk/gwas/studies/GCST90565439.
(4)IEU OpenGWAS (SNP-level PheWAS): https://opengwas.io/.
(5)FinnGen Release 12 (SNP-level PheWAS): https://r12.finngen.fi/.
(6)AstraZeneca PheWAS Portal (gene-level PheWAS): https://www.azphewas.com/.
The main MR results of this study are shown in Supplementary Table 1. The summary statistics of 1M-scBloodNL can be accessed through the resource pages above; for individual-level genotype or single-cell raw data, they should be obtained according to the application and authorization procedures of the data providers. This study did not obtain or analyze such restricted individual-level data. All other data were cited and analyzed in accordance with the terms of use of the respective sources.

## Data Availability Statement

This study performed secondary analysis using only publicly available summary statistics or public query results provided by existing studies. The data resources used and their access addresses are as follows (access date: November 4, 2026):

1. OneK1K sc-eQTL (discovery phase): https://onek1k.org/.
2. 1M-scBloodNL sc-eQTL (replication phase): https://eqtlgen.org/sc/datasets/1m-scbloodnl.html.
3. AD GWAS (GCST90565439, outcome data): https://www.ebi.ac.uk/gwas/studies/GCST90565439.
4. IEU OpenGWAS (SNP-level PheWAS): https://opengwas.io/.
5. FinnGen Release 12 (SNP-level PheWAS): https://r12.finngen.fi/.
6. AstraZeneca PheWAS Portal (gene-level PheWAS): https://www.azphewas.com/.

The main MR results of this study are shown in Supplementary Table 1. The summary statistics of 1M-scBloodNL can be accessed through the resource pages above; for individual-level genotype or single-cell raw data, they should be obtained according to the application and authorization procedures of the data providers. This study did not obtain or analyze such restricted individual-level data. All other data were cited and analyzed in accordance with the terms of use of the respective sources.

## Author Contributions

Youwen Si was responsible for study design, data analysis, interpretation of results, organization of figures and tables, and manuscript writing and revision; Youwei Si was responsible for searching, screening, and organizing the relevant literature, and assisted with submission and manuscript formatting. All authors read and approved the final manuscript.

